# Calibrating trust in AI-assisted pituitary surgery

**DOI:** 10.64898/2026.06.02.26354735

**Authors:** George Hudson, Danyal Z Khan, Feras Fayez, Sanchita Bhatia, Sophia Bano, Enrico Costanza, Ann Blandford, Danail Stoyanov, Peter McCulloch, Hani J Marcus, wider collaborators

**Author notes:** Wider collaborator list can be found in Supplementary materials (wider collaborators). **Corresponding author:** Dr George Hudson.

## Abstract

**Background:** Endoscopic endonasal transsphenoidal surgery (EETS) requires navigation around neurocritical anatomy. Today, artificial intelligence clinical decision support systems (AI-CDSSs) can orientate surgeons, but clinician trust in AI remains unclear, limiting safe deployment. This study evaluates how modifiable design affects trust and performance in a real-world pituitary surgery AI-CDSS.

**Method:** Online, 70 clinicians with pituitary surgery experience were randomised evenly to a Basic or Enhanced AI-CDSS which outline the sella on EETS operative video. The Enhanced group additionally received explanation of the model and previous publications, alongside confidence labels depicting outline reliability. Both groups annotated the sella on six video clips, first alone then with the optional AI-CDSS. Clips were ordered by declining AI performance, except for the final clip. Self-reported trust was measured using a 1-7 scale after each annotation, and performance was the DICE overlap between user annotations and the ground truth. Comparisons used Mann-Whitney U and permutation analysis.

**Results:** Sixty-four participants (91%) finished the exercise (31 Basic, 33 Enhanced). When AI performed best, median trust was 5.00 in both arms (U=559, p=.521). However, when AI performed worst, trust was significantly lower for the Enhanced group (3.00 vs 3.67, U=668, p=.035), sustained in the final clip (3.67 vs 4.33 U=687, p=.019). User performance improved with the AI-CDSS, but with no significant difference between the groups on the best or worst AI performing clips. Nevertheless, for the best AI, senior clinicians had higher median performance in the Enhanced group (0.95 vs 0.90, U=75, p=.066). There was also less dispersion in the Enhanced group when AI was inaccurate (IQR: 0.07 vs 0.21, p=.004).

**Conclusion:** Interface design can improve trust calibration in a surgical AI-CDSS and may increment performance in seniors when AI is accurate, and consistency when AI is inaccurate. In future, these features may form important safety checks during translation to the operating room.

## Introduction

Pituitary gland tumours are inherently challenging lesions to resect owing to their important neighbouring anatomy. Endoscopic endonasal transsphenoidal surgery (EETS) provides one possible corridor to facilitate safe resection^1^. However, the significant learning curve of EETS, combined with the risk of damaging neurovascular structures during dissection, means that intraoperative clinical decision-making can be challenging^2,3^. Traditional clinical decision support systems (CDSSs) such as intraoperative ultrasound or MRI disrupt the surgical workflow and can be challenging to interpret. Consequently, there is a need for alternative systems to identify these structures more seamlessly^4^.

Artificial intelligence (AI) has emerged as a new way to help clinicians make precise and useful actions^5^. In pituitary surgery, recent advances have produced a computer vision AI-CDSS that can analyse operative endoscopic video and recognise anatomy in real-time^6^. Yet the high stakes as these tools transition to clinical settings can interfere with the trust-building process and hinder implementation^7^. Trust is a complex construct but can be defined as the attitude that an agent will help achieve an individual’s goals in a situation charactered by uncertainty and vulnerability^8^. Trust which is appropriate to performance level improves the performance of the human-AI team in unreliable situations and promotes safe use of AI^9^. In the context of high-risk pituitary surgery where identification of vulnerable structures can be challenging, too little trust risks disorientation, while too much could push a surgeon into mission-critical errors.

Trust is therefore highlighted as a priority by the DECIDE-AI guidelines for early clinical AI studies which recognise that trust is not static - it may begin as a dispositional trust based on user characteristics, give way to situational trust in a particular task, and over time develop into learned trust^10,11^. Prior work in surgical AI has suggested that the relationship between trust and AI benefit may also depend on interface design: studies using AI-generated anatomical outlines have demonstrated greater performance benefit for juniors^6^, whereas other work has found greater benefit from AI among experts, potentially due to the inclusion of confidence metrics that may have enhanced trust calibration^12^.

Thus, whilst confidence labels and user guidance are important design features, their impact on the dynamic nature of human-AI trust in surgery remains unknown. The current study addresses this gap and investigates how modifiable AI design features influence the development of user trust in a real-world pituitary surgery AI-CDSS.

## Methods

### Study Objectives and Ethics

This was a pre-clinical IDEAL stage 0 study evaluating trust in an AI-CDSS which outlines sella anatomy during EETS. This study was performed according to the DECIDE-AI guidelines which emphasise the importance of studying the users of AI in early clinical work^10^. The primary objective was to establish the effect of AI explanations and confidence labels on trust in the AI-CDSS. This study received ethics approval and was registered with the ID UCLIC-2024-009. All participants provided informed consent.

### Participants and eligibility

The sample size of 70 participants was targeted pragmatically, in line with other studies which have measured trust dynamics or the impact AI confidence indicators^13,14^. Participants were recruited through professional networks across UK pituitary surgery centres. Participants were neurosurgery doctors or medical students over the age of 18 with pituitary surgery experience or equivalent. Those who could not communicate in English, provide informed consent, or those triggered by movement effects or flashing lights were excluded.

### Study Design

This was a randomised trial conducted online assessing the impact of design interface on participant trust using superiority analysis.

Participants were invited to annotate the sella on real operative videos with and without the help of an AI-CDSS which outlined the sella. Operative videos were selected from a secure database of endoscopic transsphenoidal pituitary surgeries held by a tertiary neurosurgery centre where all patients had given consent. A total of six, short five-second clips were chosen from six different operations: for three of these videos, the anatomy was relatively clear lending to superior AI performance, whereas the other three had challenging anatomy or obscuring features leading to poorer AI performance. The study was therefore not a representation of real-world AI accuracy but a simulation of variability.

Participants annotated the final frame of these clips by outlining the sella using an online platform. The platform was constructed in JavaScript and deployed with Vercel so that participants could access the platform using a website link. An introductory video tutorial was provided to familiarise participants with the platform. After annotating the six clips first without any AI to allow them to overcome the learning curve with the online platform, participants repeated their annotations, this time using an AI-CDSS showing an overlay of the sella in the clip which would then vanish. Participants could make the AI overlay reappear by toggling it on or off.

The first five clips were presented to participants in an order of declining AI accuracy and confidence to capture the ability of participants to safely recognise when AI began to perform poorly. On the sixth clip a video was presented with intermediate AI accuracy and confidence to evaluate reversibility (**figure 1**).

**Figure 1:**
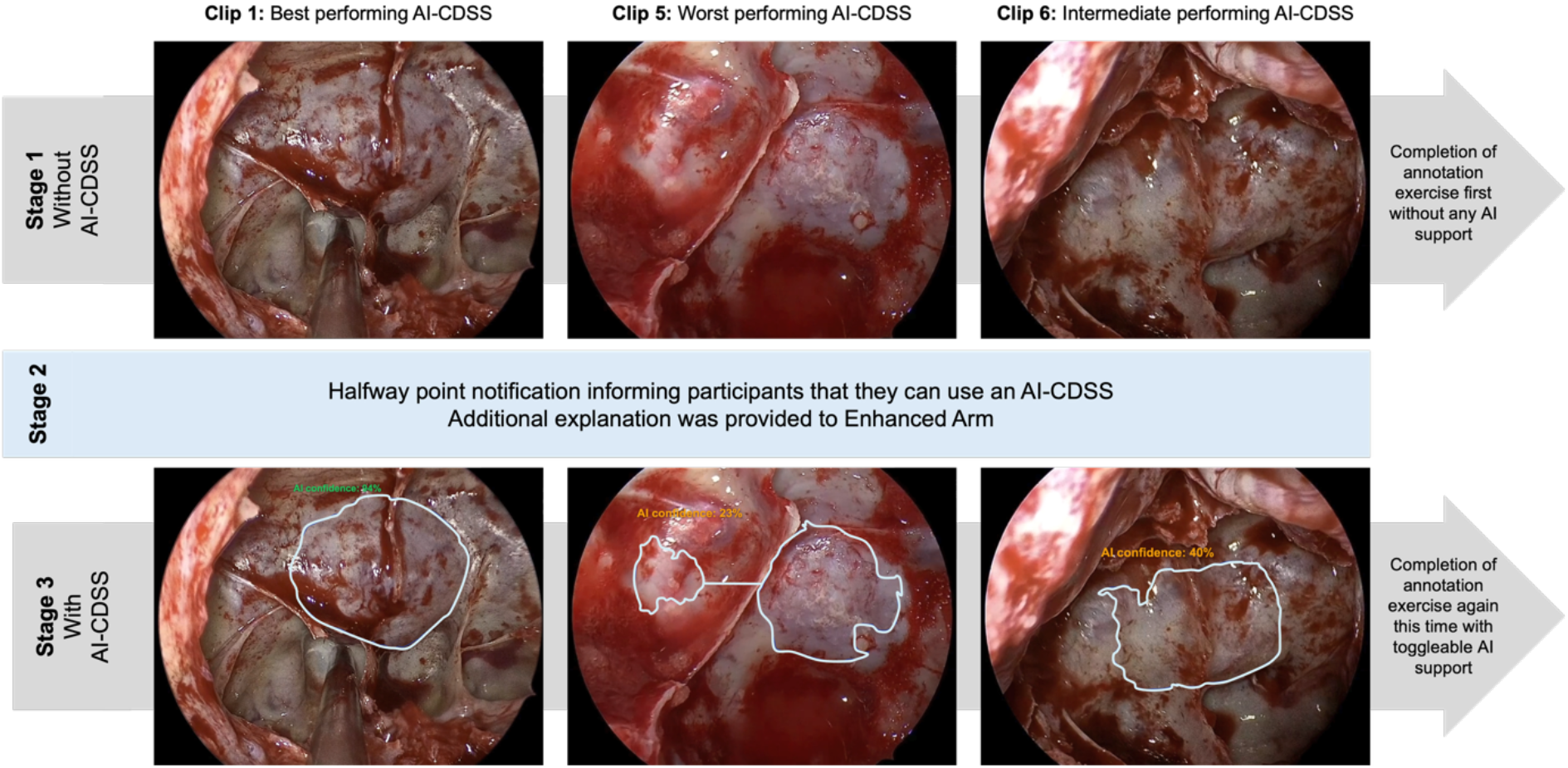
Study flow. Participants first completed six annotation exercises without an AI-CDSS and subsequently completed the same set with a toggleable AI-CDSS. Those in the Enhanced arm received additional explanation about the architecture and past performance of the AI, as well as confidence metrics on each overlay as shown in the images. Those in the Basic AI-CDSS arm were simply told they had access to AI and received overlays without any confidence readings.

### AI-CDSS

The AI used to provide the overlays is a real-world AI system which recognises anatomy in real-time^6,15,16^. Briefly, this system relies on a supervised deep neural network trained on 10 images from the sella phase of 64 anonymised videos of EETS. The ground truth was determined by three neurosurgical trainees and two consultant neurosurgeons, resulting in a multi-round expert consensus process. The training dataset and AI backbone was identical to that described in previously published work^6^.

### Intervention and randomisation procedure

The AI model was run on the six test clips, and the outputs were used to form the AI-CDSS. For the AI-assisted section of the study, participants were randomised 1:1 to either a ‘Basic’ AI-CDSS or an ‘Enhanced’ AI-CDSS. Randomisation was performed using permutated random block design with blocks of sizes 6-10 and adaptive randomisation to achieve balancing. The allocation sequence was automatically generated and implemented by the online platform and hidden from researchers. Participants were not blinded but were not aware of the existence of other versions of the AI-CDSS, data analysts were not blinded. The Enhanced AI-CDSS provided participants with further information about the AI model’s real-world structure and previously published results, alongside confidence metrics on the AI-CDSS toggle overlay depicting its reliability (**supplementary figure 1, link: https://ai-trust-app.vercel.app/**). As expected, three of the overlays were considered good quality by experienced clinicians (HM and DK) and were given fabricated confidence indicators of >70% displayed in green text. Three of the overlays were poorer quality and were given fabricated confidence indicators <70% displayed in amber text. The confidence indicators scaled with the AI vs ground truth DICE coefficients. The Basic AI-CDSS group was provided with the AI outlines with no supplementation.

### Outcome measurement

The primary outcome was trust in the AI-CDSS. Baseline characteristics known to influence dispositional trust such as participant age^17^ and biological sex^18^ were captured. Experience level was also captured in alignment with previous studies^6^. Participant personality was assessed using the Ten Item Personality Inventory (TIPI) questionnaire which has been has been validated and used in similar trust-in-AI settings^19^. Finally ‘propensity-to-trust’ was measured using a validated tool described elsewhere^20^.

The Trust In Automation Scale (TIAS) is a well-established survey which seeks to quantify such trust^21^, and recently a shortened version of this scale (STIAS) has been validated in the context of an AI assistant recommendation^22^. The STIAS metric is a mean average of three questions scored on a 1-7 Likert scale: “I am confident in the AI assistant”; “The AI assistant is reliable”; “I can trust the AI assistant”. In the current study, the STIAS survey was presented after the participant completed each annotation with the AI and was non-skippable. Performance was recorded as the DICE coefficient between the participant annotation and the expert consensus ground truth. Time spent with the AI toggled on as a proportion of overall annotation time was recorded for exploratory analysis.

### Statistics

Standard statistical analysis was performed using Python 3.11.2 through VS Code and the alpha for statistical significance was 0.05, with all tests two-sided for significance. Normally distributed data were presented as a mean and standard deviation, or median and interquartile range if not normally distributed, evaluated visually. Due to strong ceiling effects, outliers, and a relatively small sample size, Mann-Whitney U tests were used as the primary test to compare the distributions for the best and worst clip to establish the sensitivity of trust and performance. Supplementary analysis in R version 4.3.3 (2024-02-29) was performed using linear mixed-effects models fitted using glmmTMB with Gaussian error structure and random intercepts for participants. Nested models were compared using likelihood ratio tests based on differences in log-likelihood. In all cases, assumptions were interrogated visually. All randomised participants were included in the analysis, and missing data was handled using list-wise deletion.

## Results

Seventy participants were recruited between October 2025 and March 2026 and randomised to either the Basic AI-CDSS (n=35, 50%) or the Enhanced AI-CDSS (n=35, 50%) (**table 1**). The cohort was predominantly male (86%) and had even recruitment of both juniors (50% pre-residency and residency) and seniors (50% post-training attendings and fellows). Of those initiating the study, 64 (91%) went on to complete it in full (Basic AI-CDSS=31, 48%; Enhanced AI-CDSS=33, 52%) - four left before completing any of the exercises and therefore were blind to their arm allocation, two left after completing the first unassisted set of videos, both in the Enhanced AI-CDSS arm. It was not possible to determine the reason for drop-out.

**Table 1:** Baseline demographics. Categorical variables are presented as frequency (%). Continuous variables are presented as mean (s.d.).

| Variable |  | Basic<br>AI-CDSS<br>(N = 35) | Explainable<br>AI-CDSS<br>(N = 35) | Total<br>(N = 70) |
| --- | --- | --- | --- | --- |
| Sex | Female | 5 (14) | 5 (14) | 10 (14) |
|  | Male | 30 (86) | 30 (86) | 60 (86) |
| Age group | 18-24 | 0 (0) | 1 (3) | 1 (1) |
|  | 25-44 | 28 (80) | 23 (66) | 51 (73) |
|  | 45-64 | 6 (17) | 11 (31) | 17 (24) |
|  | 65+ | 1 (3) | 0 (0) | 1 (1) |
| Grade | Pre-residency | 6 (17) | 2 (6) | 8 (11) |
|  | Residency | 14 (40) | 13 (37) | 27 (39) |
|  | Post-training* | 15 (43) | 20 (57) | 35 (50) |
| Personality trait<br>(scale 1-7) | Extraversion | 4.29 (1.47) | 4.30 (1.55) | 4.29 (1.50) |
|  | Agreeableness | 4.56 (1.06) | 4.80 (0.93) | 4.68 (0.99) |
|  | Conscientiousness | 5.66 (1.15) | 5.77 (0.88) | 5.71 (1.02) |
|  | Stability | 5.36 (1.09) | 5.41 (1.18) | 5.39 (1.13) |
|  | Openness | 5.34 (1.22) | 5.26 (0.89) | 5.30 (1.06) |
| Propensity-to-trust<br>(scale 1-5) |  | 3.39 (0.69) | 3.07 (0.65) | 3.23 (0.69) |

### AI Performance

The performance of the AI as measured by the DICE coefficient against the ground truth was 0.95, 0.93, 0.91, 0.85, 0.68 and 0.86 for clips one to six respectively. The confidence metrics prescribed to each clip for the Enhanced arm accounted for the accuracy of the AI and the underlying volatility of the AI output and were 94%, 81%, 75%, 55%, 23% and 40% respectively.

### Dynamic Trust

Trust was measured on a scale 1-7 after each clip. Trust was highest in clip one, with the Enhanced AI-CDSS (median=5.00, IQR=4.67-5.67) generating similar trust scores to the Basic AI-CDSS (median=5.00, IQR=5.00-5.83, U=559, p=.521). Both arms experienced the lowest trust values for clip five where the AI performed the worst, however the trust distribution for the Enhanced AI-CDSS (median=3.00, IQR=2.00-3.67) was significantly lower compared to the Basic AI-CDSS (median=3.67, IQR=3.00-4.67, U=668, p=.035) with a small-to-moderate effect size (r=.26) (**figure 2**) (**supplementary table 1**).

**Figure 2:**
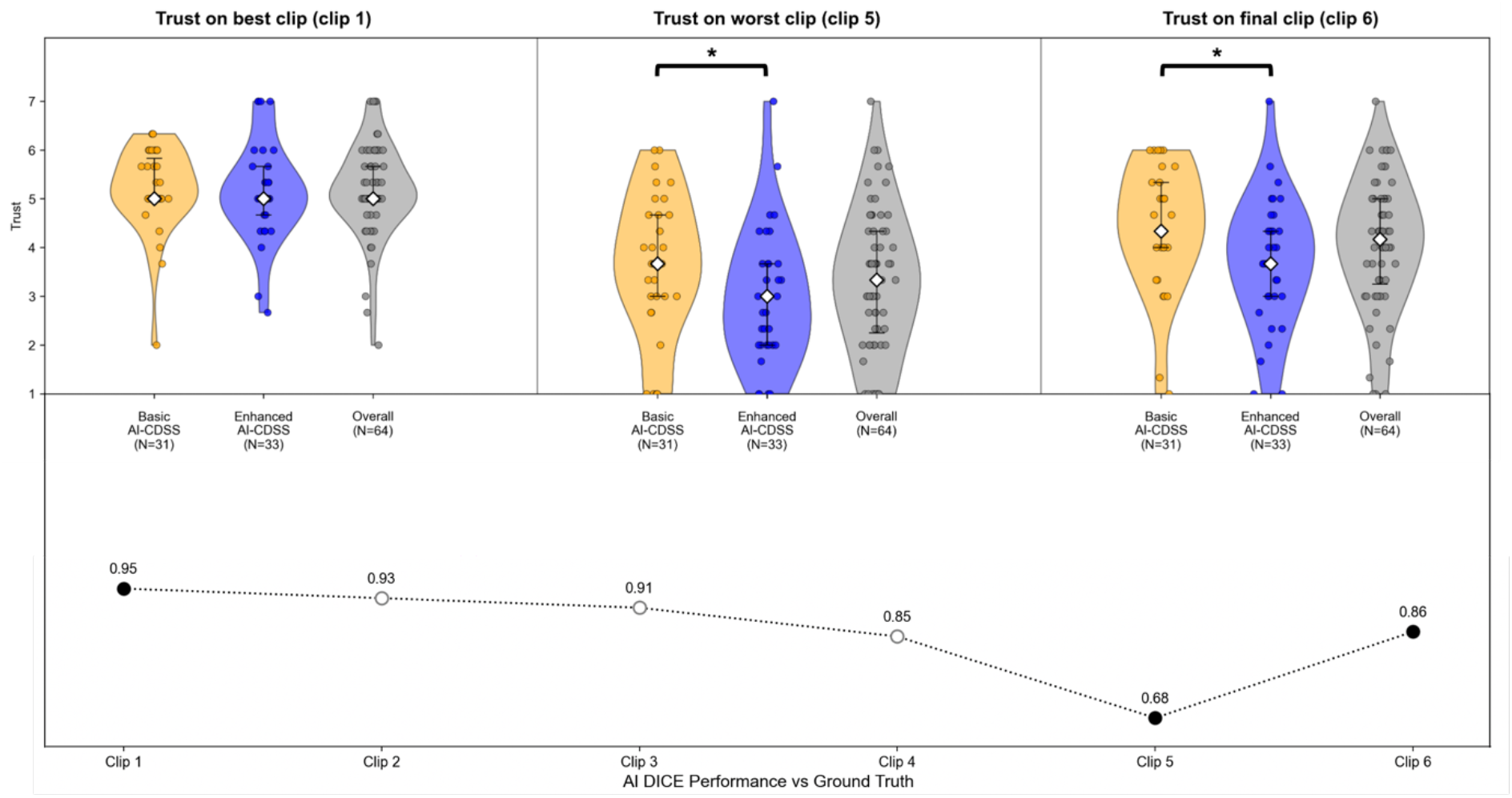
Violin plot demonstrating participant-reported trust in the best and worst clips by AI performance and the final clip. AI performance by clip number is shown in the panel below. Individual points represent unique participants; the white diamond represents within-group median and error bars represent interquartile range. * = p<.05 using Mann-Whitney U tests.

**Figure 3:**
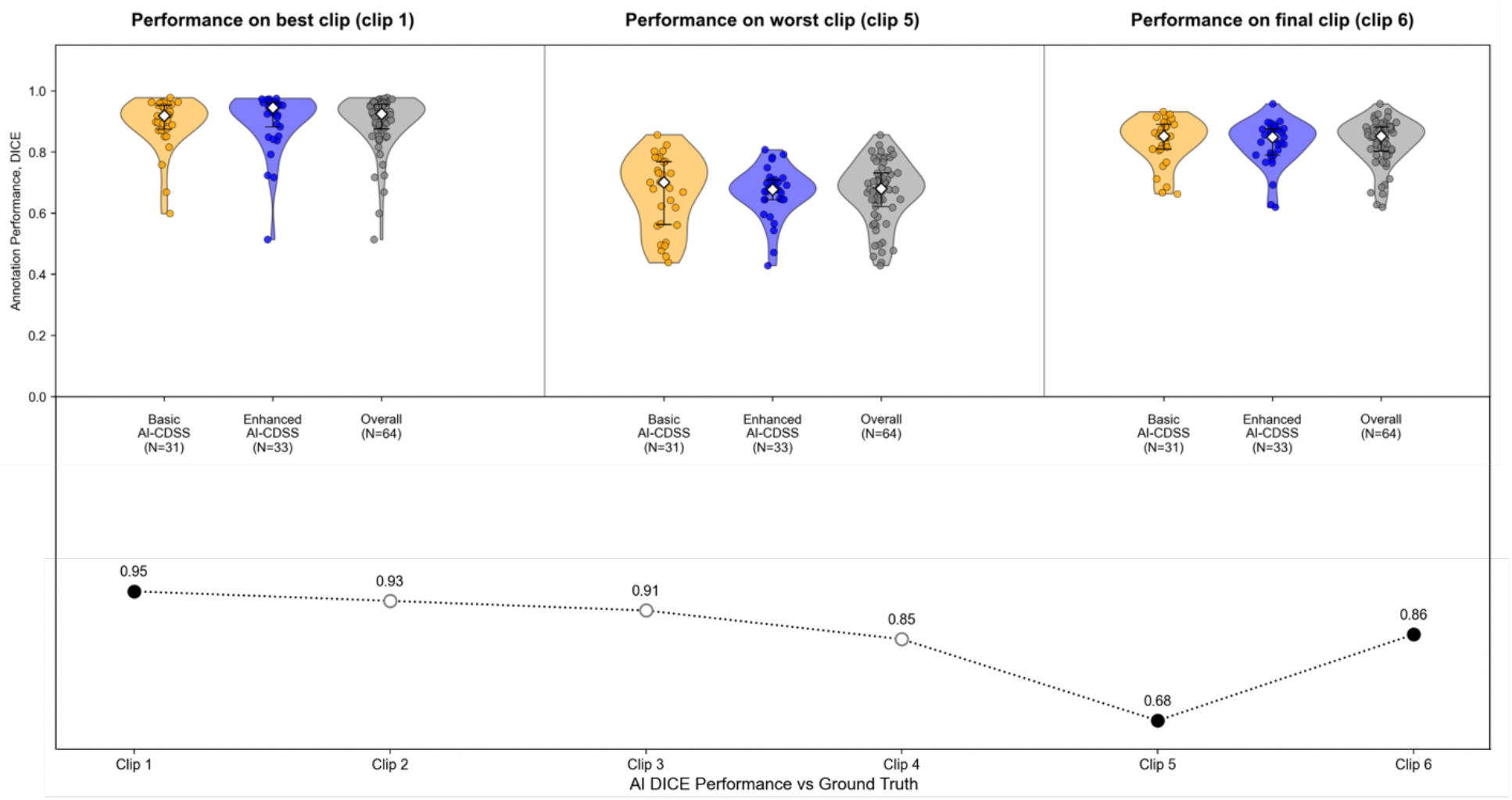
Violin plot demonstrating participant performance, measured as the DICE score between participant annotation and the ground truth, in the best and worst clips by AI performance and the final clip. AI performance by clip number is shown in the panel below. Individual points represent unique participants; the white diamond represents within-group median and error bars represent interquartile range. No significant differences were found using Mann-Whitney U tests.

For the final clip, the AI performed better, and overall median trust improved from 3.33 (IQR=2.25-4.33) to 4.17 (IQR=3.25-5.00). There remained however a significant difference in the trust distributions between the Enhanced AI-CDSS (median=3.67, IQR=3.00-4.33) and the Basic AI-CDSS (median=4.33, IQR=4.00-5.33, U=687, p=.019) again with a small-to-moderate effect size (r=.29). Trust across all clips is presented in **supplementary figure 2**.

A robustness analysis using the full repeated measures dataset showed that inferior AI performance was the dominant driver of lower trust (β = −0.56, 95% CI [−0.69, −0.43], p <.001) (**supplementary table 2**), but inclusion of study arm and its interaction with AI performance significantly improved model fit and AIC relative to a model with AI performance alone (Δχ^2^(2)=7.45, p =.024) (**supplementary table 3**).

### Performance

Performance was taken as the DICE coefficient between the participants’ annotations and the expert-derived ground truth and varied across the clips (**figure 4**). Median performance across the unassisted videos was 0.83 (IQR=0.75-0.86) which improved to 0.85 with AI assistance (IQR=0.81-0.87, W=384, p<.001) (**supplementary table 3**). Exploratory investigation of participant time spent with the AI can be found in **supplementary figure 3**.

Performance with the AI-CDSS was highest for clip one (median=0.92, IQR=0.88-0.96) and lowest for clip five (median=0.68, IQR=0.62-0.73), recovering in clip six (median=0.85, IQR=0.80-0.88). There was no significant difference in the performance distributions between the two arms of the study in any of these clips (U=437, p=.320; U=546, p=.648; U=562, p=.502 respectively).

For clip one, a trend was noted towards improved performance amongst seniors in the Enhanced AI-CDSS arm (Basic AI-CDSS median=0.90, IQR=0.87-0.93; Enhanced AI-CDSS median=0.95, IQR=0.94-0.96; U=75, p=.066) with a moderate effect size (r=.33) (**supplementary table 3**). Meanwhile, clip five demonstrated reduced variability in the Enhanced AI-CDSS group, with a narrower interquartile range (IQR=0.64-0.71) compared with the Basic AI-CDSS group (IQR=0.56-0.77). A permutation test using 50,000 permutations demonstrated that this difference in dispersion was statistically significant (p=.004).

## Discussion

This study is the first to evaluate the impact of interface design on trust in a real-world surgical AI-CDSS, tracking user trust after a series of six annotation tasks on clips taken from EETS. Trust was strongly affected by AI performance and deteriorated to a greater extent with the Enhanced AI-CDSS compared to the Basic AI-CDSS as AI performance worsened, suggesting better trust calibration. Human-AI team performance did not differ between the two groups, and both showed improved DICE performance when AI performed well, as expected. There was however a trend towards improved performance with the Enhanced AI-CDSS in seniors when AI performed well and reduced variability overall in the Enhanced AI-CDSS group when AI performed poorly.

### When AI performs well: the impact of interface on user performance

In the Enhanced AI-CDSS group, providing explanation about the model’s previous results and mechanics did not affect user trust in clip one where AI performance was the highest. The reason for this was likely multifactorial. Automation bias, whereby individuals attribute disproportionate trust to a machine they assume to be competent, has been demonstrated previously in clinical scenarios where AI delineates anatomy^23^. This may have augmented self-reported trust in the Basic AI-CDSS, obscuring the effect of the intervention. Additionally, the explanation provided may have been insufficient to impact trust. Explainable AI (XAI) often depicts not only how the model works but how the AI decision is affected by a change in circumstances^24^. Comparatively the AI-CDSS deployed here is a black-box system, limiting the ability to provide these higher levels of explanation.

Despite similar trust scores, senior clinicians demonstrated a trend towards better DICE performance with the Enhanced AI-CDSS compared to the Basic AI-CDSS for clip one. Generally, junior clinicians show greater reliance on the AI-CDSS^6^, and senior clinicians may be quicker to dismiss even a trustworthy AI due to their own higher subject matter expertise^11^. In previous research on cruise-control systems, priming users with more engaging explanation of the automation increased exploratory use^25^. Thus, whilst interface design choices did not affect initial trust, they may have encouraged senior clinicians to engage more with AI. Moreover, previous work has found that senior clinicians can benefit more from accurate AI since they may ‘cherry-pick’ the best parts of AI^12^. In this way, improvement in engagement may explain the differential benefit of experts in the Enhanced AI-CDSS group when AI performed well.

### When AI performs poorly: the impact of interface on trust calibration

As the experiment progressed, the AI-CDSS became less reliable, reaching a nadir at clip five, and trust likewise declined, particularly for the Enhanced AI-CDSS group. This agrees with prior research where confidence labels modify the trust curve and cause deterioration of trust when at 80% or below^13^. This finding is clinically meaningful to pituitary surgeons as anatomy during EETS can be highly variable, incurring risk to surrounding neurovascular structures if misunderstood^26^.

However, there was no significant difference in DICE performance between the two arms at clip five. This disconnect between trust and outcomes has been shown elsewhere. In work evaluating an automated X-ray screener, only perceptual accuracy was correlated with task performance, not trust sensitivity^27^. Similarly, another study showed that AI with confidence labels was able to improve trust calibration but did not affect the overall accuracy of the human-AI team^14^.

Since the learning curve of EETS can span hundreds of cases^3^, it is possible that early trust dynamics, which reflect calculus-based trust^28^, may not have stabilised to yet shape the decision-making processes in surgery which evolve over longer periods of time. Furthermore, the confidence labels used were not boundary-specific. Thus, even though users of the Enhanced AI-CDSS did not trust the AI, they were unable to perform better than their baseline ability. A single confidence label may therefore encourage conservative annotations but fail to allow the fine-tuning required to boost average performance. This is also true for the wider AI and automation literature: in cases of uncertainty, providing more actionable explanations has helped human-AI combined performance^29^, meanwhile unsuitable explanations have been linked to reduced user accuracy^30^. More conservative annotations might also explain the improved consistency Enhanced AI-CDSS users in clip five, guarding against particularly poor performance which would be dangerous in surgery.

### Limitations

The work presented here has several limitations. First, the study had a relatively small sample size, prone to sampling bias. Regarding study design, trust sensitivity was not known *a priori* so clips were deterministically ordered based on AI performance to improve effect resolution. However, this meant that it was challenging to statistically model the data without multi-collinearity, precluding analysis of hysteresis and of wider clip-based factors affecting trust. Finally, a different relationship between trust and performance could emerge as these tools translate into clinical practice, especially since the current study was online in absence of real risk which governs trust modelling^11^, and user environment was not controlled which is known to affect trust^31^.

## Conclusions

Despite these limitations, this work remains clinically meaningful. AI navigation systems are increasingly gaining momentum in surgery: alongside our pituitary AI-CDSS, recent work has developed systems capable of segmenting critical views in general surgery^32^, vascular structures in hepatic surgery^33^, nerves in colorectal surgery^34^ and solid organs in urological surgery^35^. However, the determinants of human-computer interaction with these surgical systems have not previously been evaluated despite being a target area of the DECIDE-AI guidelines^10^.

The current study tackles this gap, highlighting how design can improve trust calibration and achieve more consistent performance for surgical navigation tools. In this way, it provides broader insights for the clinical translation of AI-CDSSs into the operating room.

## Data Availability

All data produced in the present study are available upon reasonable request to the authors

