## Supplementary data for "Calibrating trust in AI-assisted pituitary surgery"

### *Collaborative Authorship

| Collaborator | Institution |
|---|---|
| Abhiney Jain | University College London, London, UK |
| Adam Williams | North Bristol NHS Trust, Bristol, UK |
| Ahmad Ozair | McGill University, Montreal, Canada |
| Ahmed Abougamil | Hull Teaching Hospitals NHS Foundation Trust, Hull, UK |
| Alessandro Paluzzi | University Hospitals Birmingham NHS Foundation Trust, Birmingham, UK |
| Alfonso Lagares | Department of Neurosurgery, Hospital Universitario 12 de Octubre, Spain |
| Alireza Shoakazemi | Queens Hospital, Romford, UK |
| Amir Rafati Fard | University of Cambridge, UK |
| Anastasios Giamouriadis | Aberdeen Royal Infirmary North of Scotland Brain & Spine Centre NHS Grampian, Aberdeen, UK |
| Andrew F. Alalade | Royal Preston Hospital, Preston, UK |
| Andrew Martin | Atkinson Morley Wing, St George's Hospital, London, UK |
| Angelos Kolias | Cambridge University Hospitals, Cambridge, UK |
| Anna Pogodina | University of Buckingham, Buckingham, UK |
| Anouk Borg | National Hospital for Neurology and Neurosurgery, London, UK |
| Arjun Adapa | Columbia University, New York, USA |
| Ashwin Kumaria | National Hospital for Neurology and Neurosurgery, London, UK |
| Attila Sarkadi | Hirslanden Zürich, Zürich, Switzerland |
| Carlos Botella-Campos | National Hospital for Neurology and Neurosurgery, London, UK |
| Caroline Hayhurst | University Hospital of Wales, Cardiff, UK |
| Cathal Hannan | The Walton Centre, Liverpool, UK |
| Daniel Prevedello | The Ohio State University, Ohio, USA |
| David Rowland | National Hospital for Neurology and Neurosurgery, London, UK |
| Elina Malsagov | Johannes Wesling Klinikum Minden Hospital, Minden, Germany |
| Feras Fayez | University College London, London, UK |
| Francesco Magni | Department of Neurosurgery, Sheffield Teaching Hospitals, Sheffield, UK |
| Gabriel Zada | University of Southern California, California, USA |
| George E. Richardson | Royal Lancaster Infirmary, Lancashire, UK |
| Georgios Tsermoulas | Queen Elizabeth Hospital, Birmingham, UK |
| Giorgio Fiore | Fondazione IRCCS Ca' Granda Ospedale Maggiore Policlinico, Milan, Italy |
| Hugo Layard Horsfall | National Hospital for Neurology and Neurosurgery, London, UK |
| Igor Paredes | Department of Neurosurgery, Hospital Universitario 12 de Octubre, Spain |
| Ivan Cabrilo | Neurosurgery Department, Neurocenter of Southern Switzerland, Ente Ospedaliero Cantonale, Lugano, Switzerland |
| Jan Erik Brigino Detran | Philippines |
| Joachim Starup Hansen | University College London, London, UK |
| Joao Paulo Almeida | Indiana University, Indiana, USA |
| Jonathan Funnell | Royal Sussex County Hospital, Brighton, UK |
| Jonathan Shapey | Kings College Hospital, London, UK |
| Juan C. Fernandez-Miranda | Stanford University, California, USA |
| Juan Casado Pellejero | Miguel Servet University Hospital, Zaragoza, Spain |
| Lauren Harris | National Hospital for Neurology and Neurosurgery, London, UK |
| Lewis O'Brien | University of Glasgow, Glasgow, UK |
| Ludovico Agostini | Fondazione Policlinico Universitario Agostino Gemelli, IRCCS, Italy |
| Mark A Hughes | University of Edinburgh, Edinburgh, UK |

| | |
|---|---|
| Munashe Veremu | National Hospital for Neurology and Neurosurgery, London, UK |
| Mykhailo Bandrivskyi | National Hospital for Neurology and Neurosurgery, London, UK |
| Nick Thomas | Kings College Hospital, London, UK |
| Nicola Newall | Barts Health NHS Trust, London, UK |
| Nigel Mendoza | Imperial College Healthcare NHS Trust, London, UK |
| Nina Yoh | Columbia University, New York, USA |
| Olivier Sluijters | North Bristol NHS Trust, Bristol, UK |
| Robert G. Briggs | University of Southern California, California, USA |
| Robert Spencer | University Hospital of Wales, Cardiff, UK |
| Rory J. Piper | University College London, London, UK |
| S. Bulent Omay | Yale University, Conneticut, USA |
| Samiul Muquit | University Hospitals Plymouth NHS Trust, Plymouth, UK |
| Sanchita Bhatia | University College London, London, UK |
| Shahzaib Ahmed | Queen's Medical Centre, Nottingham, UK |
| Sherry Liu | University College London, London, UK |
| Siddhant Kumar | Oxford |
| Silvia Vázquez Sufuentes | Hospital Universitario Vall d´Hebrón, Barcelona, Spain |
| Simon Williams | National Hospital for Neurology and Neurosurgery, London, UK |
| Syed Mansoor Shah | Queen Elizabeth Hospital, Birmingham, UK |
| Theofanis Giannis | National Hospital for Neurology and Neurosurgery, London, UK |
| Thomas Baron | National Hospital for Neurology and Neurosurgery, London, UK |
| Tjasa Zaletel | University College London, London, UK |

**Supplementary table 1:** Median trust scores (interquartile range) by clip number. Juniors are defined as medical students, pre-residents and residents, whilst seniors are defined as those who have completed their formal training and are senior clinical fellows or consultants.

| **Outcome** | **Subgroup** | **Basic AI-CDSS (N = 31)** | **Enhanced AI-CDSS (N = 33)** | **Total (N = 64)** | **U** | **p** | **r** |
|---|---|---|---|---|---|---|---|
| Trust on best AI clip (clip 1) | Overall | 5.00 (5.00–5.83) | 5.00 (4.67–5.67) | 5.00 (5.00–5.67) | 558 | .521 | .08 |
| | Juniors | 5.00 (5.00–5.67) | 5.00 (4.42–5.33) | 5.00 (4.92–5.67) | 154 | .278 | .19 |
| | Seniors | 5.00 (5.00–6.00) | 5.00 (5.00–6.00) | 5.00 (5.00–6.00) | 124 | 1.000 | .00 |
| Trust on worst AI clip (clip 5) | Overall | 3.67 (3.00–4.67) | 3.00 (2.00–3.67) | 3.33 (2.25–4.33) | 668 | **.035** | .26 |
| | Juniors | 3.83 (3.00–4.92) | 3.50 (2.58–4.33) | 3.67 (3.00–4.67) | 144 | .492 | .12 |
| | Seniors | 3.33 (2.67–4.67) | 2.33 (2.00–3.00) | 2.83 (2.00–3.67) | 166 | .109 | .28 |
| Trust on final clip (clip 6) | Overall | 4.33 (4.00–5.33) | 3.67 (3.00–4.33) | 4.17 (3.25–5.00) | 686 | **.019** | .29 |
| | Juniors | 4.67 (4.00–5.58) | 4.00 (3.08–4.83) | 4.33 (3.33–5.33) | 165 | .141 | .26 |
| | Seniors | 4.33 (3.33–5.00) | 3.67 (3.00–4.33) | 4.00 (3.00–4.67) | 161 | .153 | .25 |

**Supplementary table 2:** Fixed effects from the full mixed-effects regression model predicting trust. Baseline group was Basic AI-CDSS at zero AI inaccuracy (i.e. 100% DICE). *AI inaccuracy represents [1 - AI DICE coefficient vs ground truth] to ease interpretability.

| Term | Estimate (β) | 95% CI | p value |
|---|---|---|---|
| Intercept | 5.32 | 5.00 to 5.64 | <.001 |
| Enhanced AI-CDSS | -0.35 | -0.79 to 0.09 | .120 |
| AI Inaccuracy* | -0.56 | -0.69 to -0.43 | <.001 |
| Enhanced AI-CDSS x AI Inaccuracy* | -0.14 | -0.32 to 0.05 | .151 |

**Supplementary table 3**: Comparison of nested linear mixed-effects models predicting trust. The full model included confidence condition and its interaction with AI inaccuracy. Models were compared using likelihood ratio tests. *AI inaccuracy represents [1 - AI DICE coefficient vs ground truth].

| Model | Predictors | AIC | logLik | $\Delta\chi^2$ (df) | p-value |
|---|---|---|---|---|---|
| Null | AI Inaccuracy* | 1142.2 | -567.11 | | |
| Full | AI Inaccuracy* + Enhanced features + Interaction | 1138.8 | -563.39 | 7.45 (2) | .024 |

**Supplementary table 4:** Performance expressed as the DICE coefficient between participant annotation and the ground truth, stratified by participant training grade. Wilcoxon Signed Rank tests are used for comparing unassisted and assisted attempts. Mann-Whitney U tests are used for comparing the Basic AI-CDSS and Explainable AI-CDSS arms. Juniors are defined as medical students, pre-residents and residents, whilst seniors are defined as those who have completed their formal training and are senior clinical fellows or consultants.

| Outcome | Subgroup | Unassisted (N=64) | AI-assisted (N=64) | W | p | Basic AI-CDSS (N=31) | Enhanced AI-CDSS (N=33) | U | p | r |
|---|---|---|---|---|---|---|---|---|---|---|
| Overall performance across all clips | Overall | 0.83 (0.75-0.86) | 0.85 (0.81-0.87) | 384 | **<.001** | 0.84 (0.80-0.88) | 0.85 (0.82-0.87) | 542 | .687 | .05 |
| | Juniors | 0.81 (0.73-0.86) | 0.83 (0.81-0.88) | 58 | **<.001** | 0.83 (0.79-0.89) | 0.83 (0.81-0.87) | 136 | .718 | .07 |
| | Seniors | 0.84 (0.80-0.86) | 0.85 (0.82-0.87) | 150 | **.032** | 0.85 (0.81-0.87) | 0.85 (0.82-0.86) | 132 | .759 | .06 |
| Performance on best AI clip (clip 1) | Overall | 0.94 (0.84-0.96) | 0.92 (0.88-0.96) | 821 | .143 | 0.92 (0.87-0.95) | 0.95 (0.88-0.96) | 437 | .320 | .13 |
| | Juniors | 0.90 (0.76-0.95) | 0.92 (0.87-0.95) | 157 | **.045** | 0.92 (0.89-0.96) | 0.92 (0.85-0.94) | 149 | .393 | .15 |
| | Seniors | 0.95 (0.92-0.96) | 0.95 (0.88-0.96) | 262 | .978 | 0.90 (0.87-0.93) | 0.95 (0.94-0.96) | 75 | .066 | .33 |
| Performance on worst AI clip (clip 5) | Overall | 0.68 (0.64-0.74) | 0.68 (0.62-0.73) | 906 | .370 | 0.70 (0.56-0.77) | 0.68 (0.64-0.71) | 546 | .648 | .06 |
| | Juniors | 0.72 (0.64-0.75) | 0.70 (0.61-0.75) | 221 | .432 | 0.73 (0.58-0.78) | 0.68 (0.64-0.71) | 158 | .231 | .21 |
| | Seniors | 0.68 (0.64-0.72) | 0.68 (0.64-0.70) | 239 | .651 | 0.68 (0.56-0.70) | 0.68 (0.65-0.70) | 108 | .565 | .11 |
| Performance on final clip (clip 6) | Overall | 0.82 (0.75-0.86) | 0.85 (0.80-0.88) | 561 | **.001** | 0.85 (0.81-0.89) | 0.85 (0.79-0.88) | 562 | .502 | .08 |
| | Juniors | 0.80 (0.72-0.87) | 0.85 (0.80-0.88) | 111 | **.003** | 0.85 (0.81-0.88) | 0.85 (0.79-0.87) | 138 | .662 | .08 |
| | Seniors | 0.83 (0.78-0.86) | 0.85 (0.81-0.88) | 173 | .091 | 0.85 (0.81-0.90) | 0.85 (0.79-0.88) | 139 | .565 | .11 |

**Supplementary figure 1:** A screenshot of the explanation of the AI tool which was provided to those in the Explainable AI-CDSS arm prior to completing annotations with the AI-CDSS.

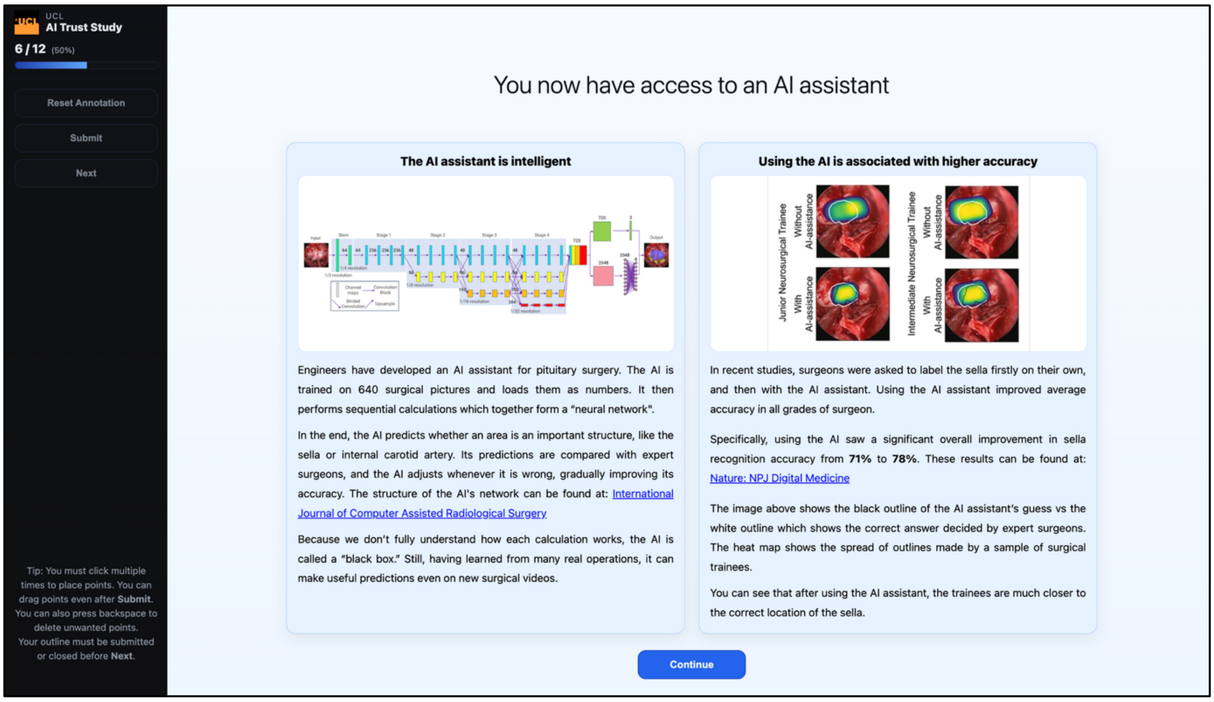


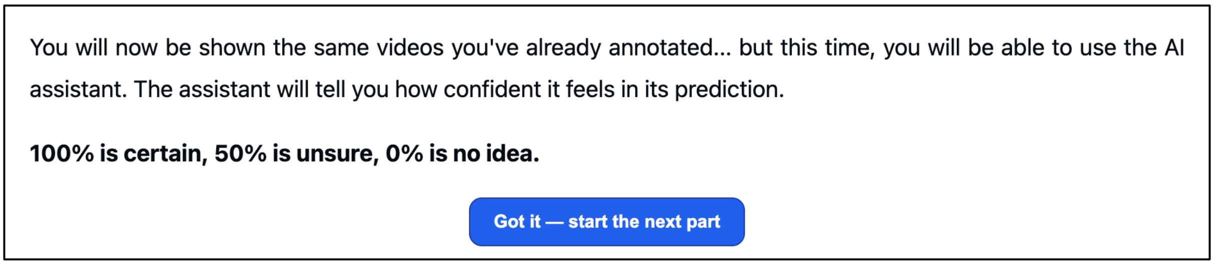

**Supplementary figure 2:** Descriptive chart demonstrating how trust changed by clip using box plots with the median line in black.

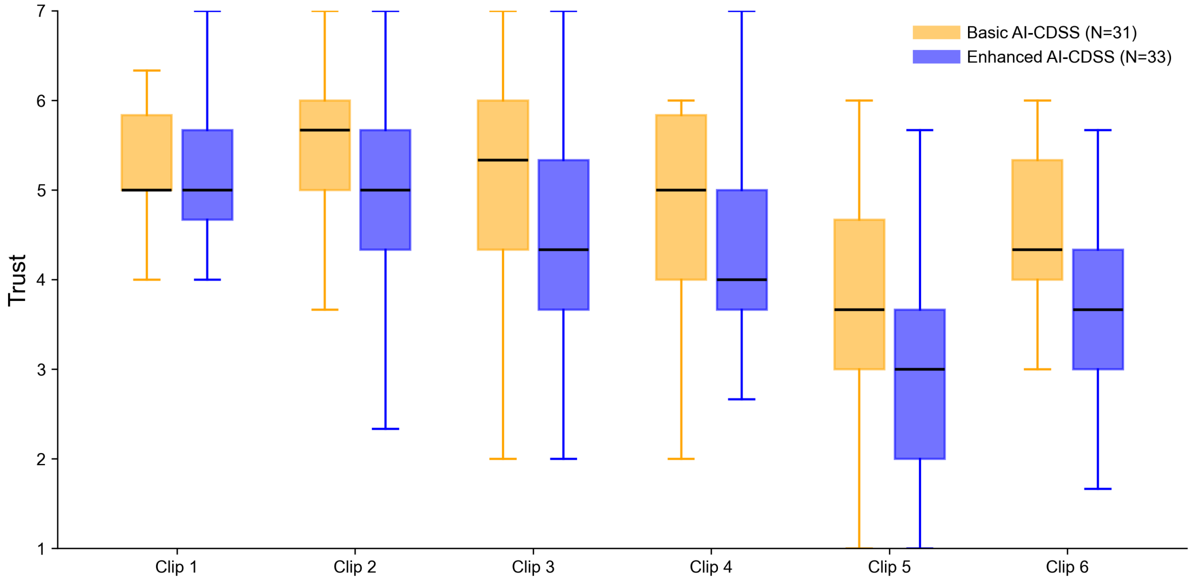

**Supplementary figure 3:** Sankey diagram demonstrating the adjustment of observation strategy throughout the clip set. Heavy use is defined as keeping the AI-CDSS toggled on greater than or equal to 50% of the total time spent on the still image. Light use is defined as keeping the AI-CDSS toggled on less than 50% of the total time spent on the still image. Persistent users kept using the same strategy throughout.

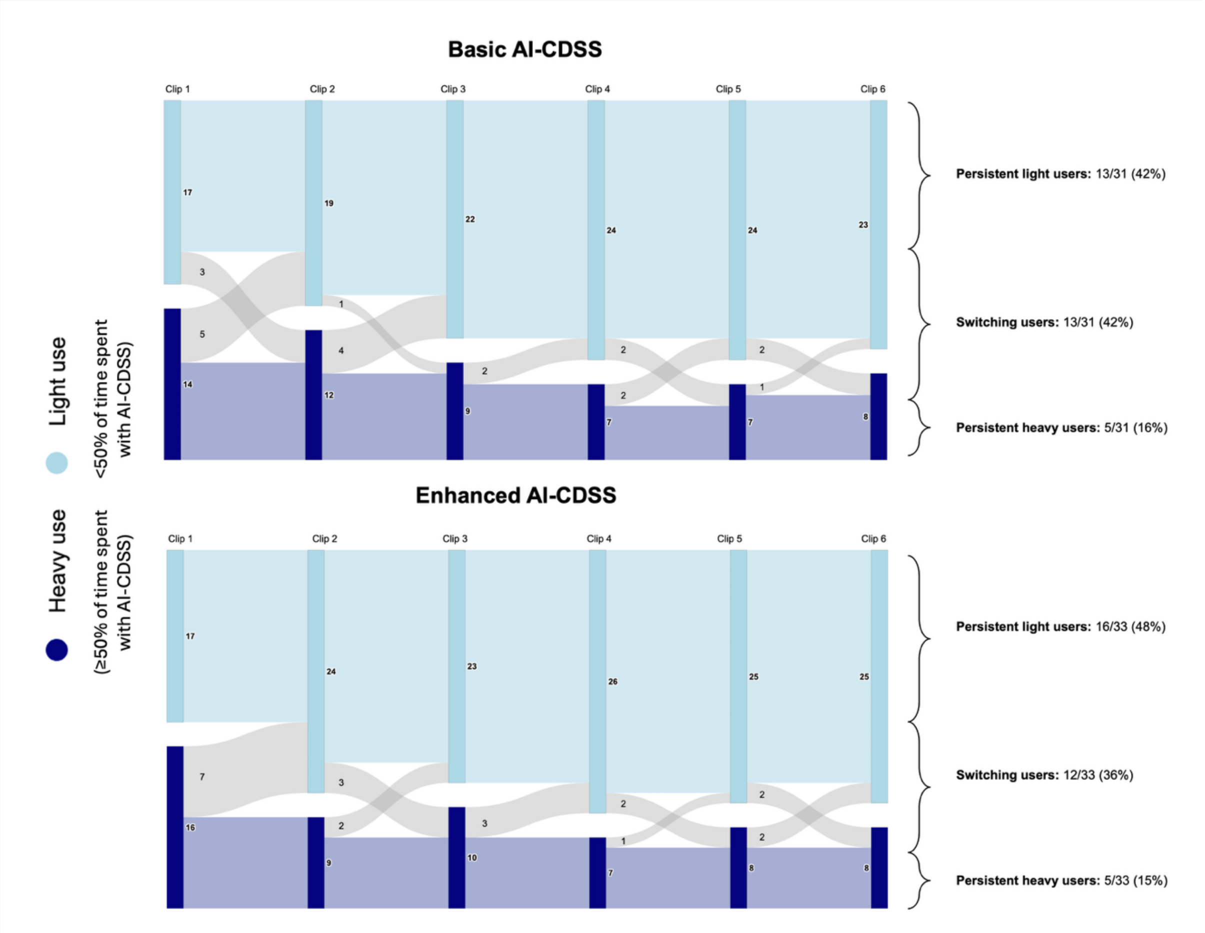
